# Use of multiple Polygenic Risk Scores for distinguishing Schizophrenia-spectrum disorder and Affective psychosis categories; the EUGEI study

**DOI:** 10.1101/2021.03.31.21254574

**Authors:** Victoria Rodriguez, Luis Alameda, Diego Quattrone, Giada Tripoli, Charlotte Gayer-Anderson, Edoardo Spinazzola, Giulia Trotta, Hannah E Jongsma, Simona Stilo, Caterina La Cascia, Laura Ferraro, Daniele La Barbera, Antonio Lasalvia, Sarah Tosato, Ilaria Tarricone, Elena Bonora, Stéphane Jamain, Jean-Paul Selten, Eva Velthorst, Lieuwe de Haan, Pierre-Michel Llorca, Manuel Arrojo, Julio Bobes, Miguel Bernardo, Celso Arango, James Kirkbride, Peter B Jones, Bart P Rutten, Alexander Richards, Pak C Sham, Michael O’Donovan, Jim Van Os, Craig Morgan, Marta Di Forti, Robin M Murray, Evangelos Vassos

**Author notes:** Joint senior author; these authors made similar contributions (Robin M Murray, Evangelos Vassos). Corresponding author details: Department of Psychosis Studies., Institute of Psychiatry, Psychology and Neuroscience., King’s College London (Denmark Hill Campus), 16 De Crespigny Park, Camberwell London SE5 8AF, United Kingdom.

## Abstract

Schizophrenia (SZ), Bipolar Disorder (BD) and Depression (D) run in families. This susceptibility is partly due to hundreds or thousands of common genetic variants, each conferring a fractional risk. The cumulative effects of the associated variants can be summarised as a polygenic risk score (PRS). Using data from the EUGEI case-control study, we aimed to test whether PRSs for three major psychiatric disorders (SZ, BD, D) and for intelligent quotient (IQ) as a neurodevelopmental proxy, can discriminate affective psychosis (AP) from schizophrenia-spectrum disorder (SSD). Participants (573 cases, 1005 controls) of european ancestry from 17 sites as part of the EUGEI study were successfully genotyped following standard quality control procedures. Using standardised PRS for SZ, BD, D, and IQ built from the latest available summary statistics, we performed simple or multinomial logistic regression models adjusted for 10 principal components for the different clinical comparisons. In case-control comparisons PRS-SZ, PRS-BD and PRS-D distributed differentially across psychotic subcategories. In case-case comparison, both PRS-SZ (OR=0.7, 95 %CI 0.53-0.92) and PRS-D (OR=1.29, 95%CI 1.05-1.6) differentiated global AP from SSD; and within AP categories, only PRS-SZ differentiated BD from psychotic depression (OR=2.38, 95%CI 1.32-4.29). Combining PRS for severe psychiatric disorders in prediction models for psychosis phenotypes can increase discriminative ability and improve our understanding of these phenotypes. Our results point towards potential usefulness of PRSs for diagnostic prediction in specific populations such as high-risk or early psychosis phases.

## Introduction

More than 100 years have passed since Kraepelin established the dichotomy of manic-depression and dementia praecox as the two fundamental pillars of psychotic illness, which still constitutes the basis of current diagnostic criteria ^1^. However, it is a matter of debate whether Schizophrenia (SCZ) and Bipolar Disorder (BD) are discrete illnesses or conditions which are part of an overall conceptual continuum ^2–4^. Given the high heritability of these disorders ^5^, genetic tools can be used to dissect possible biological differences between these diagnostic categories.

Genome Wide Association Studies (GWAS) have shown that, as with other psychiatric conditions, many hundreds or thousands of common alleles influence susceptibility to SCZ and BD ^6,7^. We can calculate individual polygenic risk scores (PRS) based on the summation of the carried risk of single nucleotide polymorphisms (SNPs) selected in a discovery GWAS according to their *p*-value, weighted by their effect size ^8,9^. GWAS analyses by the Psychiatric Genomics Consortium (PGC) have estimated liability–based SNP-heritability for SCZ, BD and Major Depressive Disorder (MDD) as about 22.2% ^10^, 18.2% ^7^, and 8.5% ^11^ respectively in case–control samples.

In line with previous family and twin studies ^12–14^, GWAS findings have also supported the notion of genetic overlap among severe mental disorders. A study from the Cross-Disorder Group of PGC ^15^ showed genetic correlation using common SNPs, of around 0.70 between SCZ and BD, 0.34 between SCZ and MDD, and 0.36 between BD and MDD.

On the other hand, some studies provide support for a link between genetic predisposition and current diagnostic categories. A study investigating diagnostic subcategories across the psychosis spectrum employing PRS for SCZ and BD (PRS-SZ and PRS-BD) ^16^ provided some validation for the existence of subcategories across the SCZ and BD continuum. In line with this, in a more recent study, PRS-SZ discriminated SCZ from BD; and within BD subtypes, between those with and without psychosis ^17^. Moreover, Markota et al. ^18^, found that PRS-SZ seemed to be more closely related with BD type I with psychotic symptoms during manic phases as compared with BD-I with psychotic symptoms during depressive episodes or presenting without psychosis. Taken together, these findings shed light on the genetic architecture of these severe mental disorders and support the discriminability potential of the polygenic score on diagnostic categories.

Despite this evidence, most studies have only tested the association between PRS-SZ and PRS-BD with their respective diagnostic categories. To the best of our knowledge, only one study has previously examined the relationship between diagnostic categories by employing three polygenic scores, specifically PRS-SZ, PRS-BD and PRS-MDD ^19^, but only examined cases within the BD spectrum. They found a PRS-SZ gradient among affective psychotic categories, with the highest association being schizoaffective followed by BD type I and BD type II.

Consistent evidence suggests that cognitive deficits can be considered a core feature for schizophrenia ^20^. It has been long accepted that subjects affected by SCZ perform worse than those with BD on a variety of cognitive domains ^21,22^, which seems to be validated by a meta-analysis showing that subjects with BD show better cognitive performance than those with SCZ ^23^. Although there remains debate over the extent to which these differences in cognition predate or follow the onset of psychosis ^24^, it suggests the hypothesis that PRS for measures of cognitive ability including intelligence may be informative for studying genetic differences between these subgroups of patients.

Given the above, the current study aims to explore the potential of joint modelling PRS from three major mental disorders (SCZ, BD, D) and intelligence quotient (IQ) for firstly, analysing the distribution of genetic load of major psychiatric disorders across the diagnostic categories under the psychosis umbrella, thus helping us understand whether current diagnoses represent different genetic subgroups; and secondly, exploring the potential use of PRSs in discriminating affective psychosis (AP) from schizophrenia-spectrum disorder (SSD). We built on a previous study from South London, where it was shown that PRS-SZ differentiated schizophrenia from other psychoses ^25^. In a time of growing interest in employing PRS as a tool for validating phenotypes or diagnosis, we aim to explore the potential of joint modelling PRS in discriminating AP from SSD, hypothesizing that PRS can be used to distinguish between diagnostic categories.

## Methods

### Sample

The present study is based on the case-control sample from the EUGEI study (EUropean Network of national schizophrenia networks studying Gene-Environment Interactions); a multisite incidence and case-control study of genetic and environmental determinants involved in the development of psychotic disorders ^26^.

The baseline sample comprises a total of 2627 participants, including 1130 patients aged 18 to 64 years who were resident within the study areas and presented to the adult psychiatric services between May 1, 2010 and April 1, 2015 in 17 sites across 6 countries: England, the Netherlands, Italy, France, Spain and Brazil. All participants provided informed, written consent. Ethical approval was provided by relevant research ethics committees in each of the study sites. All data was stored anonymously.

Cases were selected if they were experiencing their first episode of psychosis (FEP) including SCZ and related psychosis, BD and Major Depression Disorder with Psychotic features (MDD-P). In addition, 1497 unaffected screened controls with no lifetime psychotic disorder were also recruited in the areas served by the services with a quota sampling approach, a non-probability sampling method in which a specific subgroup is chosen in order to represent the local population. Further information about the methodology of the study is available on the EU-GEI website (www.eu-gei.eu/) and can be found in previous publications ^26–29^.

One of the problems when using current PRS is the limited predictive power in multi-ethnic samples as they have derived from mostly European samples ^30^. This has been shown in a previous study on FEP patients ^25^, where PRS_SZ had much lower predictive power in African ancestry population. Given the wide variance across ancestral groups, for the scope of the present study we constrained the sample to those categorised as of European ancestry based on a Principal Component Analysis (details provided in *Supplementary Material*). Characteristics of the final sample are summarised in **Table 1**.

**Table 5.1.** Sociodemographic of white subsample (n=1659), case-control comparisons.

| DESCRIPTIVE AT BASELINE | Number (%)/<br>Mean(SD) |  | Statistics |  | Number (%)/<br>Mean(SD) | Statistics |  |
| --- | --- | --- | --- | --- | --- | --- | --- |
|  | Control<br>n= 1005 | Schizophreni<br>a-spectrum<br>disorder<br>n= 409 | Tests (df) | p value | Affective<br>psychosis<br>n= 164 | Tests (df) | p value |
| Gender |  |  | X<sup>2</sup> (1)=50.54 | <0.001 |  | X<sup>2</sup>(1)=0.67 | 0.413 |
| Male | 474 (47.2) | 278 (68) |  |  | 83 (50.6) |  |  |
| Female | 531 (52.8) | 131 (32) |  |  | 81 (49.4) |  |  |
| Age (years) | 36.9 (13) | 31.63 (10.92) | Z=7.21 | <0.001 | 32.84 (11.56) | Z=-3.76 | <0.001 |
| EVER USED CANNABIS |  |  | X<sup>2</sup> (1)=40.26 | <0.001 |  | X<sup>2</sup>(1)=15.9 | <0.001 |
| No | 528 (53) | 136 (34.2) |  |  | 58 (36) |  |  |
| Yes | 469 (47) | 262 (65.8) |  |  | 103 (64) |  |  |
| EDUCATION LEVEL |  |  | X<sup>2</sup> (2)=81.22 | <0.001 |  | X<sup>2</sup> (2)=51.64 | <0.001 |
| No qualification | 40 (4) | 65 (16.1) |  |  | 25 (15.3) |  |  |
| School education | 416 (41.5) | 197 (48.6) |  |  | 87 (53.4) |  |  |
| Tertiary education | 546 (54.5) | 143 (35.3) |  | <0.001 | 51 (31.3) |  | <0.001 |
| YEARS IN EDUCATION | 14.69 (4.19) | 12.94 (4.12) | Z=7.07 |  | 12.58 (3.84) | Z=5.92 |  |
| SOCIAL FUNCTIONING |  |  |  |  |  |  |  |
| Employment status |  |  | X<sup>2</sup> (1)=25.26 | <0.001 |  | X<sup>2</sup> (1)=0.48 | 0.487 |
| Employed | 615 (61.6) | 141 (45.5) |  |  | 79 (58.5) |  |  |
| Unemployed | 383 (38.4) | 169 (54.5) |  |  | 56 (41.5) |  | 0.001 |
| Marital status |  |  | X<sup>2</sup> (1)=126.23 | <0.001 |  | X<sup>2</sup> (1)=11.42 |  |
| Steady relationship | 626 (62.4) | 105 (28.3) |  |  | 74 (48.1) |  |  |
| No relationship | 378 (37.7) | 266 (71.7) |  |  | 80 (52) |  | 0.001 |
| Living arrangements |  |  | X<sup>2</sup> (1)=96.98 | <0.001 |  | X<sup>2</sup> (1)=11.85 |  |
| Independent living | 683 (68.5) | 119 (37.5) |  |  | 73 (53.7) |  |  |
| No independent living | 314 (31.5) | 198 (62.5) |  |  | 63 (46.3) |  |  |

SD: standard deviation; df: degrees of freedom

### Measures

#### Diagnosis

We used DSM-IV diagnosis ^31^ from interviews and mental health records utilizing the Operational Criteria Checklist (OPCRIT) at baseline ^32^ by centrally trained investigators, whose reliability was assessed throughout the study (κ = 0.7). These diagnoses were grouped into: schizophrenia spectrum disorders group (SSD - codes 295.1-295.9 and 297.1-298.9 -) or affective psychosis group (herein called AP-patients diagnosed with codes 296-296.9), which was later stratified into BD (codes 296.0-296.06 and 296.4-296.89) and MDD with psychotic features (MDD-P – codes 296.2-296.36-). For those subjects with missing information for DSM-IV output from OPCRIT, we reconverted ICD-10 diagnosis (n=5) into DSM-IV codes; leaving eventually diagnostic data for 12 cases missing. Those who did not meet criteria from OPCRIT (i.e. undefined diagnosis) were not grouped into either of the groups (n=52) and were excluded from further analyses.

#### Genotyping and Polygenic risk scores building

DNA from blood tests or saliva sample were obtained from the majority of participants at baseline (73.6% of cases and 78.5% of controls), with no sociodemographic differences observed with those without genetic data except for minor age differences (please refer to the *Supplementary material* section1.7). All DNA data collected was genotyped at the Cardiff University Institute of Psychological Medicine and Clinical Neurology, using a custom Illumina HumanCoreExome-24 BeadChip genotyping array covering 570,038 genetic variants; and quality control was performed locally (details provided in *Supplementary material*).

In order to control for population stratification, a Principal Component Analysis generating 10 principal components (PC) was run on pruned variants. After quality control of genetic and clinical data, and selection of individuals of European ancestry (details provided in *Supplementary material*), the genetic analyses included 573 cases (409 SSD, 74 BD and 90 MDD-P patients) and 1005 controls.

The measure of the aggregate genetic load is based on polygenic risk score, which is an individual quantitative risk factor calculated from the weighted summation of the odds ratios of carried risk alleles taken from a discovery sample. It is represented by the following equation ^33^:

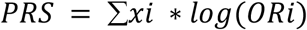

where *x* is the number of risk alleles of each included variant (*i*) and OR the respective odds ratio. To build the PRSs, results from the latest available GWAS which did not include the current EUGEI sample, were used as discovery samples. In the case of SCZ and BD, these were derived from the last mega-analyses of the PGC ^7,10^. Depression PRS was built from a GWAS combining PGC, 23andMe and UK Biobank ^7,10,34^. Finally, we further included the recently developed PRS for IQ ^35^. All PRS were built using PRSice software ^36^, and the selected p-value threshold of 0.05 for SNP inclusion was chosen across the phenotypes on the basis of the published literature explaining the most variance in case-control analysis ^11,34,35,37^. Each PRS was standardized to a mean of zero and standard deviation of 1 ^38^.

### Statistical analyses

#### Descriptive statistics

Normality of all variables was assessed computing Shapiro-Wilk normality test. The comparisons between cases and controls and between AP and SSD cases were made using chi-square, t-test or Wilcoxon-Mann-Whitney tests when appropriate. Effect sizes were calculated for all the statistical tests using Cohen’s d for t-test and Cramer’s V (Fc) for chi-square. When Mann-Whitney test was used, effect sizes were calculated from z values.

#### Association analyses

We first analysed PRSs association with broad clinical groups (SSD, AP) compared with controls; and in a second step in a case-only analysis we measured discrimination ability of PRSs between AP categories (BD and MDD-P) and SSD as reference group. For this, we built a series of multinomial or simple logistic regression models in which we included the three disorder PRSs (PRS-SZ, PRS-BD, PRS-D) plus PRS-IQ as independent variables while controlling for population stratification using as confounders the 10 PC and each sample site. Due to the inclusion of the four PRSs in the models, we adjusted the significance level as per Bonferroni’s correction ^39^, with a new established significance level at p<0.0125. Results will be presented in OR, 95% confidence intervals (CI) and p-value. We conducted power calculation analyses utilising the R-package AVENGEME ^8^, which allows power calculation for PRS analyses. We calculated the required SNP-h^2^ or fix covariance in our target sample to obtain 80% of power on each regression model and per each PRS (SZ, BD and D).

#### Fitness of model for SSD and AP discrimination

As a secondary analysis we explored goodness of fit of data of the joint use of PRSs. We built a series of logistic regression models to test discriminability between AP and SSD in which we sequentially added one PRS at a time in order to identify those PRS adding significant value to the discriminability between the clinical groups by comparing models through likelihood ratio test (see *Supplementary material* for more details).

## Results

### Socio-demographics

Socio-demographics of the case-control sample are shown in **Table 1**, comparing SSD (n=409) and AP (n=164) with controls (n=1005) separately. Compared with controls, patients were younger (mean age of 31.6, SD=10.91 and 32.84, SD=11.56 in SSD and AP respectively; 36.9, SD=13 in controls); and a greater proportion of patients with SSD were men (68% vs 47%). Both SSD and AP were less likely to have received tertiary education and consequently reported fewer total years of education than controls (around over 12.5 years in cases and around 14.7 years for controls). Generally, cases were more likely not to be in a relationship and not to live independently. More SSD patients were unemployed, but no differences between AP and controls were found.

### PRS distribution in different clinical subgroups (model 1)

The first multinomial logistic regression model showed that higher scores on both PRS-SZ and PRS-BD were associated with SSD (OR=1.87, 95%CI 1.57-2.2, p<0.001 and OR=1.34, 95%CI 1.15-1.57, p<0.001 respectively), whereas positive associations with AP were found for PRS-BD and PRS-D (OR=1.35, 95%CI 1.09-1.67, p=0.006 and OR=1.37, 95%CI 1.14-1.64, p=0.001 respectively) compared with controls. These effects are shown in **Figure 1** with additional details given in *Supplementary Material* (***eTable4***).

**Figure 1.**
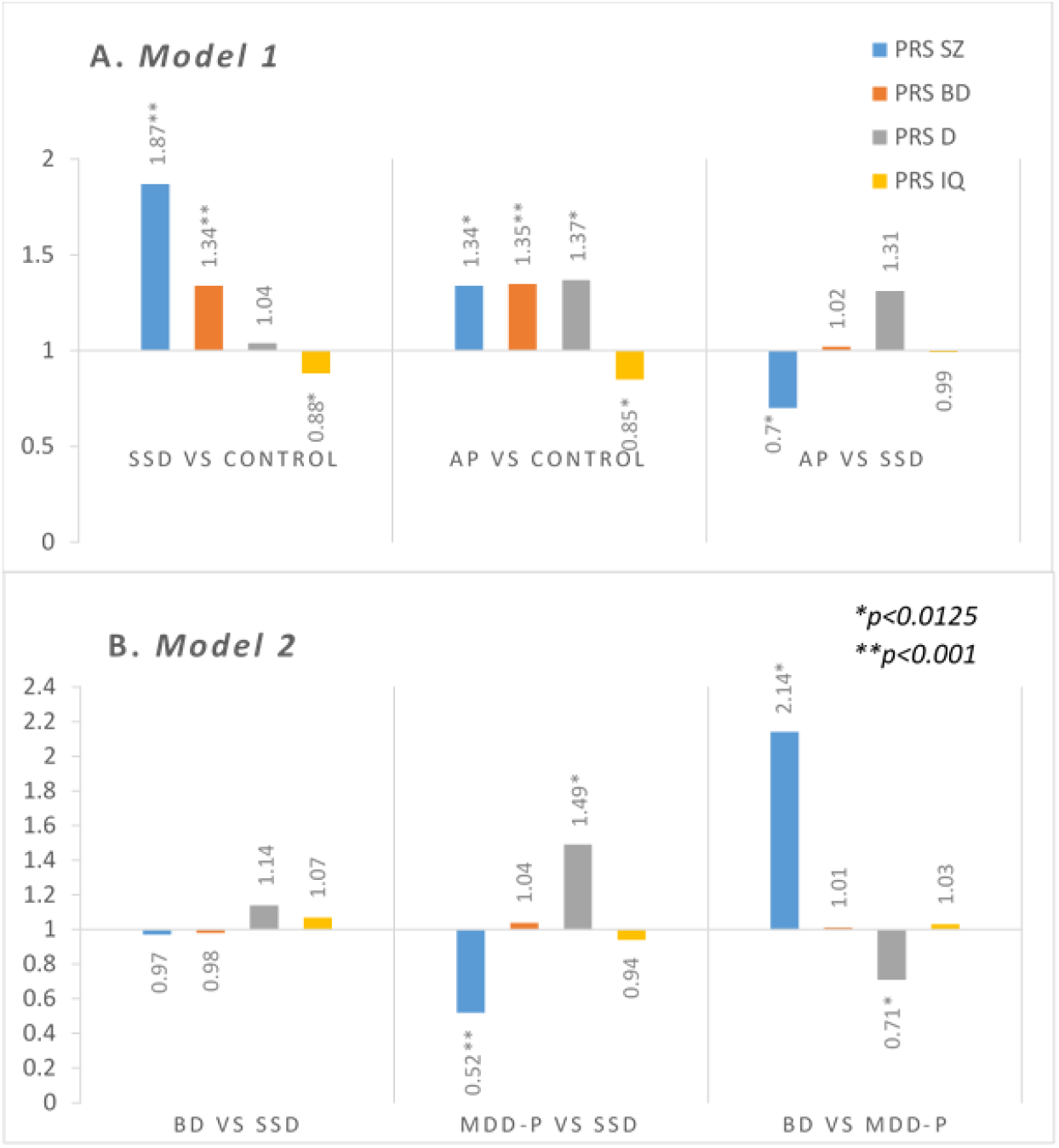
PRS performance for identifying clinical subgroups and categories based on DSM-IV OPCRIT. Results of OR from joint model with all PRSs, adjusted by 10PCs and site. SZ: schizophrenia; BD: bipolar disorder; D: depression; IQ: intelligence quotient; SSD: schizophrenia-spectrum disorder; AP: affective psychosis; MDD-P: psychotic depression. *p<0.0125 **p<0.001

In the direct comparison between AP and SSD, both PRS-SZ and PRS-D were significantly associated with these diagnoses but in opposite directions. Whereas PRS-D (OR=1.31, 95%CI 1.06-1.61, p=0.011) was associated with increased risk of AP compared with SSD, the opposite was observed for PRS-SZ (OR=0.7, 95%CI 0.54-0.92, p=0.010). Hence, individuals with high PRS-SZ and low PRS-D have more chances of receiving diagnosis of SSD, while low PRS-SZ and high PRS-D increases the chances of AP (**Figure 2**).

**Figure 2.**
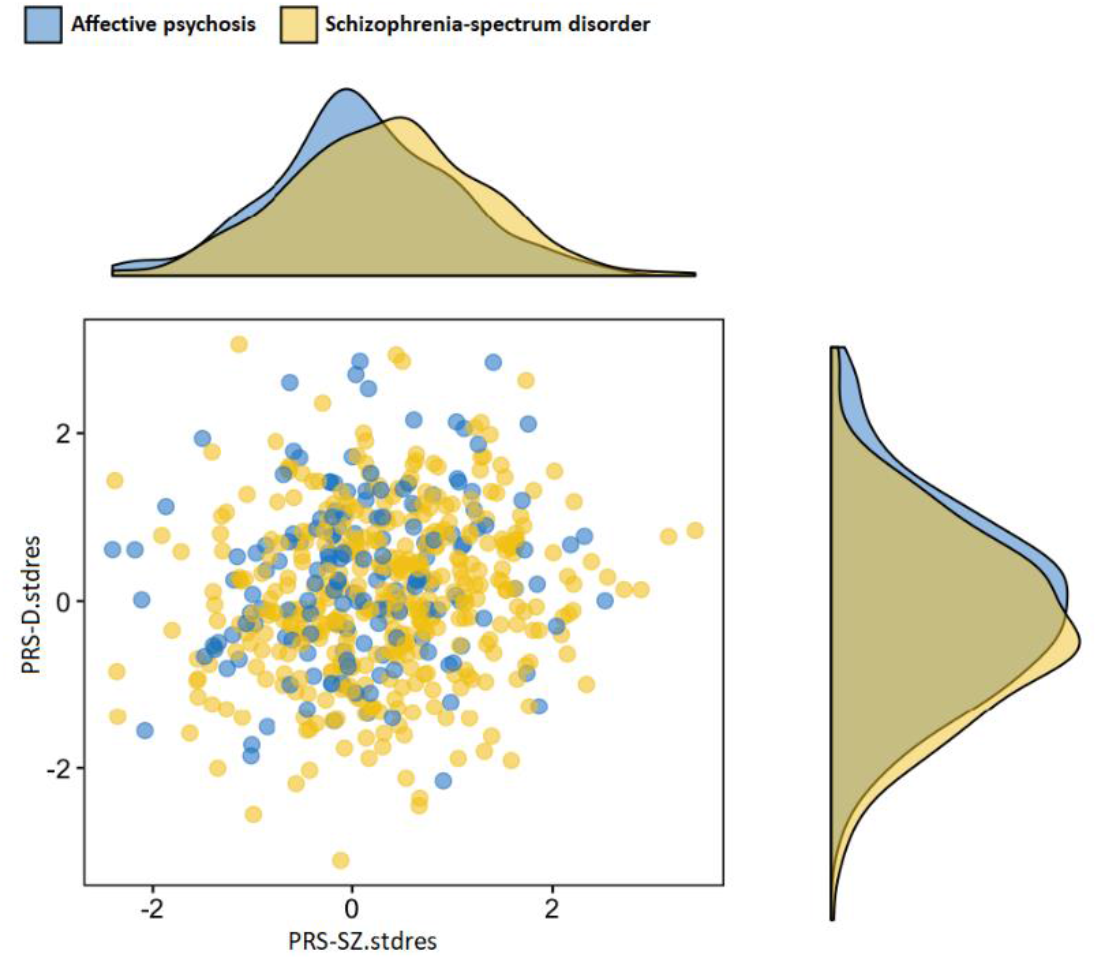
PRS-SZ and PRS-D distribution in cases with SSD and AP diagnosis. Scatterplot and density distributions of PRS-SZ and PRS-D in AP and SSD. Residuals of polygenic scores converted into z-score after adjustment for principal components and sites. Higher PRS-SZ increases the chances of SSD, while higher PRS-D increases the chances on affective psychosis

### PRS distribution between diagnostic categories within psychosis (model 2)

In model 2 we tested whether PRSs could differentiate individual diagnostic categories included in AP (BD and MDD-P) from the broad group of SSD. As shown in **Figure 1 (B)**, no PRS was able to distinguish BD when compared with SSD. Nonetheless, the patterns for SSD and MDD-P diagnoses followed those observed above for SSD and broader AP comparisons. Thus, SSD and MDD-P diagnoses were differentiated by both PRS-SZ (OR=0.52, 95%CI 0.37-0.74, p=0.011) and PRS-D (OR=1.49, 95%CI 1.14-1.94, p=0.003) in the opposite direction. Further details are given in *Supplementary Material* (***eTable5***), When running simple logistic regression for discriminability between BD and MDD-P, only PRS-SZ could discriminate people diagnosed with BD from those diagnosed with MDD-P (OR=2.14, 95%CI 1.23-3.74, p=0.007) showing a positive association with the former.

### Fitting the model optimising PRS for SSD and AP discrimination

In order to test which combination of PRSs better differentiated SSD and AP as our main outcome, we built a series of regression models sequentially including the four PRSs variables, once at a time. The best fitting data as per likelihood ratio test was by adding PRS-SZ and PRS-D to the model (Δχ^2^(1) = 6.74, p=0.0094) when compared with a model using only PRS-SZ. No further addition of PRS-BD or PRS-IQ improved the discrimination between clinical categories. Further details are provided in *Supplementary Material* (***eFigure4***)

## Discussion

To the best of our knowledge, this is the largest multisite international case-control study to examine joint polygenic associations with specific diagnostic categories in FEP patients. Our results provide evidence to support an inverse gradient of PRS-SZ and PRS-D across diagnostic categories in the psychosis spectrum, as illustrated in **Figure 3**; while they also show a discriminability potential to distinguish SSD from AP, especially from MDD-P. No PRS was able to distinguish BD from SCZ in this sample, while PRS-SZ was the only factor which distinguished BD from MDD-P. Moreover, we found that combining PRS for different disorders improves the prediction model for psychosis-related phenotypes while increasing our understanding of these phenotypes.

**Figure 3.**
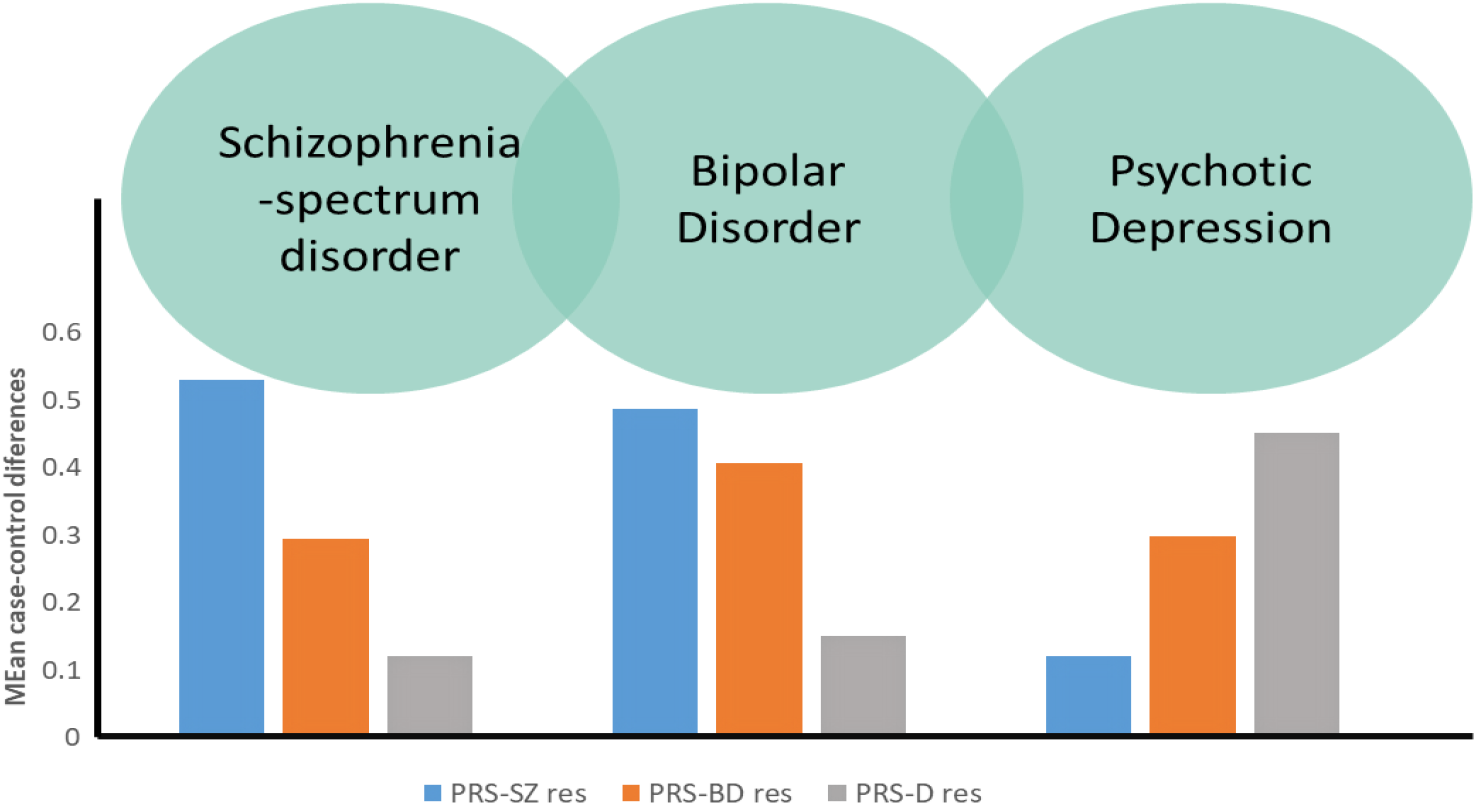
Visual representation of PRSs distribution across diagnosis categories. Conceptual multidimensional distribution of SNPs for Schizophrenia, Bipolar Disorder and Depression across clinical groups. Based on mean case-control differences, using control as reference of Standardised Residuals of PRS for SZ, BD and D adjusted by 10PC and site.

### Interpretation of findings and comparison with other studies

The observed PRS-SZ associations which followed a gradient from schizophrenia-spectrum disorder to affective diagnosis categories (SSD>BD>MDD-P), are in line with the notion of a psychosis continuum across psychosis diagnostic categories and the observed genetic overlap between disorders ^13^. Other studies have previously shown a similar PRS-SZ gradient (SZ>BD type I>BD type II) ^17,19^. However, PRS-SZ could not differentiate MDD-P from controls in our study. In a recent study, PRS-SZ seems to be specially associated with those presenting psychotic features in the mania phase when compared with the depressive pole ^18^, which could explain our lack of association with MDD-P.

Previous research showed evidence of PRS-D case-control discriminability for depression ^11^. Moreover, PRS-D failed to identify diagnostic subtypes in some case-only comparisons ^19^, but seemed to be significantly associated with schizoaffective disorder depressed subtype when compared with schizophrenia ^40^. In our study, PRS-D differentiated MDD-P from both controls and SSD, showing similar effect sizes as PRS-SZ. This may be due to the increased variance which was explained when selecting more severe patients with MDD ^41^ – only with psychotic features in our case -; the use of more powerful PRS-D built from PGC plus Biobank ^34^; or that MDD-P may be phenomenologically different to MDD without psychosis.

In relation to our second aim (ie. whether we could use PRSs in order to distinguish between affective vs schizophrenia spectrum disorder subgroups), both PRS-SZ and PRS-D differentiated global AP from SSD, and the subtype of MDD-P against SSD. Nonetheless, when trying to differentiate the categories of BD and SSD, all PRSs failed to differentiate between them. This may indicate the large genetic correlation between the two disorders, that may only be present to a lesser extent in depressive patients with psychotic features. Indeed, PRS-SZ was also able to distinguish BD from MDD-P, supporting the notion of lower common genetic liability for schizophrenia in those suffering with psychotic depression than in those with bipolar disorder.

These results shed new light on the existence of yet unclear and blurred genetic boundaries between current diagnosis categories. Beyond the evidence of a gradient for risk of psychosis associated with PRS-SZ from SSD to the AP group, we could also observe an inverse gradient in the case of PRS-D. This allows the conceptualization of a model in which the genetic vulnerability of psychotic disorders is distributed across a multidimensional continuum with SSD at one end, BD in the middle and MDD-P at the other extreme (**Figure 2**). Among these groups, only the categories in the extremes were able to be differentiated by current polygenic scores. Further studies with larger samples or when the predictive power by PRSs increase, will allow further discrimination between categories, for example between SCZ and BP or between BP and MDD-P.

We failed to observe differences in PRS-IQ distribution, although it should be noted the effect sizes are almost identical across clinical groups. Among AP, BD has been more widely compared with SCZ as the paradigm disorder within SSD. We know from previous studies that patients with BD tend to present less cognitive impairment than those with SCZ ^3,42^, but this difference seems to be less clear between individuals with SCZ and BD patients with a history of psychotic symptoms ^43^. Indeed, and in line with this, PRS-IQ showed no statistically significant differences within the case-only comparisons. However, the lack of discriminability potential of PRS-IQ would also be expected under the consideration that some cognitive changes are due to factors associated with the prodromal phase, the onset of the disorder or its treatment, rather than purely being neurodevelopmental, which is yet to be established.

### Strengths and limitations

These results should be interpreted in the context of some limitations. First, the number of patients with MDD-P and BD was relatively small which could have led to low power in analyses comparing these groups and possibly contributing to the lack of association between those categories on most PRS variables. Post-hoc power calculations of the performed comparisons showed 80% power with an estimated SNP-h^2^ in our sample lower than SNP-h^2^ of training sample for PRS-SZ, which suggest enough power for all comparisons except BD vs MDD-P comparison. Regarding PRS-BD and PRS-D, our study had 80% power to detect an association if the correlation between genetic effect of BD and depression and our BD and MDD-P phenotypes were of around 26-48% and 14-24% respectively for the highest and least powered comparisons (more detail information in *Supplementary Material*). With FEP samples a limitation to consider is the changeability of diagnoses. As shown in some studies, shifts in diagnoses occur with a predominant direction from affective psychosis to SSD in a frequency of around 14-29% after two years ^44,45^. Furthermore, comparisons between models are limited by the different discriminative power of each PRS (PRS-SZ is currently more powerful than PRS-BD and PRS-D). These models are expected to improve as bigger discovery samples are available for the affective psychotic categories. Finally, all analyses were performed in the people of European ancestry population, which limits the generalisability of the findings in other populations. However, the fact that this is a multicentre well-characterised sample of FEP, allows it to have generalisability within Caucasian European populations.

### Conclusions

Overall, this study provides support for the presence of a genetic psychosis continuum (shown by the ability of PRS-SZ to differentiate most case groups from controls following a gradient across categories). Nonetheless, we also observed genetic differences between clinical categories, with schizophrenia spectrum disorders at one end and psychotic depression at the other when looking at genetic loading for SCZ and Depression. This study also shows that combining PRSs for different disorders in a prediction model of psychosis related phenotypes improve our prediction models while contribute to our understanding of these phenotypes. Despite not yet clinically applicable at individual level, this study points towards the potential usefulness as a research tool in specific populations such as high-risk or early psychosis phases, where it may help to suggest different therapeutic approaches (i.e antidepressant versus antipsychotic) or to anticipate prognosis. However, further work is needed to explore if PRS have synergistic effects with environmental exposures before combining all the risk factors into a single prediction model.

## Data Availability

Additional data is provided in the supplementary files

## Acknowledgements

EU-GEI is the acronym of the project “European network of National Schizophrenia Networks Studying Gene-Environment Interactions”. The research leading to these results has received funding from the European Community’s Seventh Framework Programme under grant agreement No. HEALTH-F2-2010-241909 (Project EU-GEI).

Victoria Rodriguez was funded by a PhD scholarship supported by Lord Leverhulme’s Charitable Trust and by the Velvet Foundation. Evangelos Vassos is funded by the National Institute for Health Research (NIHR) Biomedical Research Centre at South London and Maudsley NHS Foundation Trust and King’s College London. The views expressed are those of the authors and not necessarily those of the NHS, the NIHR or the Department of Health and Social Care. Celso Arango was supported by the Spanish Ministry of Science and Innovation. Instituto de Salud Carlos III (SAM16PE07CP1, PI16/02012, PI19/024), co-financed by ERDF Funds from the European Commission, “A way of making Europe”, CIBERSAM. Madrid Regional Government (B2017/BMD-3740 AGES-CM-2), Fundación Familia Alonso and Fundación Alicia Koplowitz. Miguel Bernardo was supported by the Ministerio de Economía y Competitividad (PI08/0208; PI11/00325; PI14/00612), Instituto de Salud Carlos III – Fondo Europeo de Desarrollo Regional. Unión Europea. Una manera de hacer Europa, Centro de Investigación Biomédica en Red de salud Mental, CIBERSAM, by the CERCA Programme / Generalitat de Catalunya AND Secretaria d’Universitats i Recerca del Departament d’Economia I Coneixement (2017SGR1355). Departament de Salut de la Generalitat de Catalunya, en la convocatoria corresponent a l’any 2017 de concessió de subvencions del Pla Estratègic de Recerca i Innovació en Salut (PERIS) 2016-2020, modalitat Projectes de recerca orientats a l’atenció primària, amb el codi d’expedient SLT006/17/00345. Miguel Bernardo is also grateful for the support of the Institut de Neurociències, Universitat de Barcelona.

