## Supplementary Material for "Use of multiple Polygenic Risk Scores for distinguishing Schizophrenia-spectrum disorder and Affective psychosis categories; the EUGEI study"

**Supplementary Online Content**

***INDEX***

- **eAppendix 1. Methods**
  - 1.1 Sociodemographics characteristics
  - 1.2 Genotyping and PRS building
  - 1.3 PCA analyses and definition of European ancestry based on PC
  - 1.4 Diagnostic subcategories
  - 1.5 Clinical diagnosis
  - 1.6 Justification of regression model and detailed regression analysis
  - 1.7 Power calculation of main analyses
  - 1.8 Representability of included sample
- **eAppendix 2. Results (eTables and eFigures)**
  - 2.1 Sociodemographics comparison with effect sizes
    - 2.1.1 eTable 2. Case-control comparison of sociodemographic in white subsample
    - 2.1.2 eTable 3. Affective vs Schizophrenia-spectrum disorder sociodemographic comparison in white subsample
  - 2.2 Case-control variance explained of employed PRSs (eFigure1)
  - 2.3 Visual representation of data
    - 2.3.1 eFigure2. Three-dimensional scatterplot of the PRS distribution in the three groups of affective, non-affective psychosis and controls
  - 2.4. Detailed results of association of different associations on Model 1 and Model 2.
    - 2.4.1 eTable 4. Model 1: Association of different PRSs (SZ, BD, D and IQ) with multicategorical DSMIV OPCRIT clinical groups adjusted with 10 PCs in white population
    - 2.4.2 eTable 5. Model 2: Association of different PRSs (SZ, BD, D and IQ) with multicategorical DSMIV OPCRIT diagnostic variable adjusted with 10 PCs in white population (using non-affective psychosis as reference)
  - 2.5 PRS performance for identifying all psychotic diagnostic categories based on OPCRIT using control as reference (eTable 6 and eFigure3).
  - 2.6 Goodness of fit of data of join model combining three major psychiatric disorder polygenic scores (SZ, BD, MDD-P) and polygenic score for intelligence (eFigure4)
- **eBibliography**

*This supplementary material has been provided by the authors to give readers additional*

*information about their work*

**SUPPLEMENTARY**

1. **eAppendix 1. Methods**

***1.1 Sociodemographics characteristics***

Socio-demographic data was collected using the Medical Research Council (MRC) Socio-demographic Schedule modified version^1^ and supplemented by clinical records. For educational level, we stratified the sample into three categories: No qualification, school education (GCSE, ‘O’ levels and ‘A’ levels equivalent) and tertiary education (vocational, college, university or professional qualification). We dichotomized employment (employed vs. unemployed), marital status (married/in a stable relationship vs. no relationship) and living arrangement (independent living vs. no independent living).

***1.2 Genotyping and PRS building***

DNA from blood tests or saliva sample was obtained from most participants at baseline (73.6% of cases and 78.5% of controls). EUGEI sample was genotyped at Cardiff University Institute of Psychological Medicine and Clinical Neurology, using custom Illumina HumanCoreExome-24 BeadChip genotyping arrays containing probes for 570038 genetic variants (Illumina, San Diego, CA). Genotype data were called using the GenomeStudio package and transferred into PLINK format for further analysis.

Quality control was conducted in PLINK v1.07^2^ or with custom Perl scripts. Variants with call rate < 98% and with Hardy-Weinberg Equilibrium p-value < 1e-6 were excluded from the dataset. After QC, 559505 variants remained. Samples with call rate < 98% were excluded from the dataset. A linkage disequilibrium pruned set of variants was calculated using the --indep-pairwise command in PLINK (maximum r2=0.25, window size=500 SNPs, window step size = 50 SNPs) and used for further analyses. Homozygosity F values were calculated using the --het command in PLINK, and outlier samples (F < -0.11 or F > 0.15) excluded. The genotypic sex of samples was calculated from X chromosome data using the --check-sex command in PLINK, and samples with different genotypic sex to their database sex excluded.

Identity-by-descent (IBD) values were calculated for the sample in PLINK. Samples with 2 or more database siblings in the database that were not supported by the genotypic data, or with 1 or more siblings among the genotyped samples according to the database but no identified genotypic siblings (defined as PI-HAT > 0.35 and < 0.65) were excluded. After visually observing clustering of errors by genotyping chip, we decided to further exclude chips with a high proportion of errors. All samples on chips with 5 or more sample exclusions due to heterozygosity or call rate (out of 12 possible samples) were excluded. All samples on chips with 4 or more sample exclusions due to sex or relative checks were also excluded, unless their identity was corroborated by concordance between database and genotype relatedness data with a sample on another chip

For constructing PRSs, clumping was performed in imputed best-guess genotypes for each dataset using PLINK (maximum r2=0.1, window size=500kb, minimum MAF=5%), and variants within regions of long-range LD around the genome (including the MHC) excluded^3^. PRS were then constructed from best-guess genotypes using PLINK at 10 different p-value thresholds (PT=1, 0.5, 0.2, 0.1, 0.05, 0.01, 1x10^-4^, 1x10^-5^, 1x10^-6^, 5x10^-8^). We used PT=0.05 for our primary analysis, as this explained the most variation in the phenotype of schizophrenia^4^, bipolar disorder^5^, depression^6^ and IQ^7^.

***1.3 PCA analyses and definition of European ancestry based on PC***

Principal components were calculated in PLINK using LD pruned variants across the whole sample. We ranked the sample in centiles based on PC1, and calculated proportion of self-reported European ethnicity in a dichotomy fashion (yes/no) on each centile. We established the cut-off point in the stacked PC1 when three groups in a row reported less than 0.5 of whiteness. We repeated the process for PC2 and used the two cut-off point as threshold in which those who fell within them were considered as European.

***1.4 Diagnostic subcategories***

Further DSM-IV OPCRIT diagnosis^8^ were grouped as follows: Schizophrenia (DSM-IV code 295.2-295.9), Schizoaffective Disorder (DSM-IV code 295.7), other psychosis (including delusional -295.1- and psychosis NOS -298.9 -), Bipolar disorder (DSM-IV codes 296-296.06 and 296.4-296.9) and psychotic depression (codes 296.2-296.36) in order to make comparisons between them and with controls.

***1.5 Clinical diagnosis***

Additionally to those diagnosis based on OPCRIT, clinical diagnoses were received at the moment of first contact by the services, which were collected as part of the NOS-DUP scale (Nottingham Onset Schedule – Duration of Untreated Psychosis - measurement version)^9^. These diagnoses were later coded according to the DSM-IV diagnoses.

Equally, these diagnoses were grouped into: affective psychosis group (patients diagnosed with codes 296-296.9) or non-affective psychosis group (codes 295.1-295.9 and 297.1-298.9). We also had missing information for 4 cases for clinical diagnosis.

***1.6 Justification of regression model and detailed regression analysis***

We built our models on multinomial logistic regression as it is used when the dependent variable is multicategorical (or *multinomial*), when there are more than two categories and these can’t be ordered in a meaninful way. This regression model assumes that: 1) each independent variable has a specific value for each observation; 2) the independent variable can’t predict perfectly the dependent variable in any case; 3) collinearity is relatively low. For using multinomial logistic regression there is no need for the independent variables to be statistically different, and after checking for multicollinearity within our independent variables using Stata command *estat vif* , overall VIF was 2.56, which falls below suggested tolerance threshold stablish on 10.0 ^10^.

Firstly, a multinomial logistic regression model was built to compare the association of SSD and AP with controls; followed by a simple logistic regression model comparing the association between SSD and AP groups. In our second multinomial logistic model, we tested how PRSs performed in differenciating BD and PD from SSD as reference group. Additionally and only included in supplementary material we built an additional model to analyse how PRSs distribute across all psychotic diagnostic categories. We included a multicategorial variable as dependent variable using control as reference group and the four PRSs as independent variable. The diagnostic categories included in this multicategorical variable were: schizophrenia (SZ), schizoaffective disorder (SAD), other psychosis (OP), Bipolar disorder (BD) and Psychotic depression (PD). Lastly, an individual multiple logistic regression model was performed to compare PRSs associations between BD and PD.

The effect size output provided by multinomial logistic regression is Relative Risk Ratio - RRR -, which should be interpreted as the Odds Ratio between each category and always the stablished reference category. For our model 1, control is the reference category, while in model 2 we stablished as reference group the SSD.

***1.7 Power calculation of main analyses***

We conducted power calculation analyses utilising the R-package AVENGEME(Dudbridge, 2013), which allows power calculation for PRS analyses. We calculated the required SNP-h2 or fix covariance in our target sample to obtain 80% of power on each regression model and per each PRS (SZ, BD and D). Whenever the estimated covariance is too low, reflecting a SNP-h2 in the target sample lower than the SNP-h2 of the training sample, it can be considered plausible, and therefore accepted the 80% power. In our case, calculated SNP-h2 were only lower for PRS-SZ. Regarding PRS-BD and PRS-D, our study had 80% power to detect an association at p-value 0.1 if the correlation between genetic effect of BD and depression and our BD and MDD-P phenotypes were of around 26-48% and 14-24% respectively for the highest and least powered comparisons. A limitation is that this procedure only allows calculating power on associations with phenotypes tested on training samples, which prevent to calculate power of those associations between PRSs with other phenotypes (i.e associations with PRS IQ, or power of PRS BD in the “SSD vs control” association). AVENGEME calculations assumed the following values:

|  | PRS SZ | PRS BD | PRS D |
| --- | --- | --- | --- |
| Number of SNP after QC on target sample | 559505 | 559505 | 559505 |
| Training sample size | 150064 | 51710 | 807553 |
| p-value threshold in training samples | 0.1 | 0.1 | 0.1 |
| Fix variance value (SNP-h) | 0.21^4^ | 0.17-0.23 (0.20)^5^ | 0.089^6^ |
| Fix null prop to value | 0.95 | 0.95 | 0.95 |
| Training trait prevalence | 0.01 | 0.015 | 0.15 |
| Training sampling factor | 0.246488 | 0.39358 | 0.305073 |
| Target trait prevalence | 0.01 | 0.015 | 0.15 |

Estimated covariance and SNP-h^2^ values for 80% of power of the different comparison for the appropriate PRSs are provided on the table below.

| Info in target sample | Sample size | Sampling factor | Estimated covariance | Estimated SNP-h^2^ | Power |
| --- | --- | --- | --- | --- | --- |
| SSD vs CONTROL  .PRS SZ | 1414 | 0.2893 | 0.12 | 0.069 | 84% |
| AP vs CONTROL  .PRS BD  .PRS D | 1169 | 0.1403  0.1403 | 0.26  0.14 | 0.338  0.22 | 78.9%  83.6% |
| AP vs SSD  .PRS SZ  .PRS BD  .PRS D | 573 | 0.7138  0.2862  0.2862 | 0.18  0.29  0.15 | 0.154  0.42  0.25 | 80.9%  80%  81% |
| BD vs CONTROL  .PRS BD | 1078 | 0.068 | 0.37 | 0.685 | 79.3% |
| MDD-P vs CONTROL  .PRS D | 1096 | 0.083 | 0.18 | 0.36 | 81.6% |
| BD vs MDD-P  .PRS BD  .PRS D | 164 | 0.4451  0.5549 | 0.48  0.24 | 2.589  0.65 | 79.8%  78.7% |

***1.8 Representability of included sample***

No differences in gender, educational level and diagnosis but only small differences on age were found between included subjects with genotype data and those without DNA information available; suggesting a good representability of the whole sample.

*1.8.1 eTable 1. Comparison of sociodemographic of included and excluded samples based on genetic availability*

| **DESCRIPTIVE AT BASELINE** | Number (%)/Mean(SD) | | Statistics |  |
| --- | --- | --- | --- | --- |
| Gender  Male  Female   Age (years), mean (SD) | Subjects with DNA  n= 2026    1079 (53.3)  947 (46.7)  34.3 (12.3) | Subjects without PRS n= 605    328 (54.2)  277 (45.8)  33 (12.3) | Tests (df)    Χ2(1)=0.172       U=-2.7 | p value    0.679       0.009 |
| EDUCATION LEVEL  No qualification  School education  Tertiary education  YEARS IN EDUCATION | 199 (9.9)  888 (44.1)  925 (46)  14.29 (6.5) | 57 (9.6)  290 (48.9)  246 (41.5)  15.52 (12.3) | Χ2(2)=4.39  U=0.216 | 0.111  0.829 |
| OPCRIT DSM IV DIAGNOSIS  Schizophrenia-spectrum disorder  Affective psychosis  Bipolar disorder  Psychotic depression | 542 (73.1)  95 (12.8)  104 (14.1) | 197 (74.3)    35 (13.2)  33 (12.5) | Χ2(2)=0.42 | 0.811 |

*SD: standard deviation; df: degrees of freedom*

2. **eAppendix 2. Results**

***2.1 Sociodemographics comparison with effect sizes***

*2.1.1 eTable 2. Case-control comparison of sociodemographic in white subsample (n=1659)*

| **DESCRIPTIVE AT BASELINE** | Number (%)/Mean(SD) | | Statistics |  |  |
| --- | --- | --- | --- | --- | --- |
| Gender  Male  Female   Age (years), mean (SD) | Cases  n= 654    412 (63)  242 (37)   31.8 (10.95) | Control n= 1005    474 (47.2)  531 (52.8)   36.9 (13) | Tests (df)    Χ2(1)=39.91       U=7.66 | Effect sizes  V=.15 (.11-.20)  r= 0.19 | p value    <0.001       <0.001 |
| EVER USED CANNABIS  No  Yes | 224 (35.3)  410 (64.7) | 528 (53)  469 (47) | Χ2(1)=48.46 | V=0.17 (0.13-0.22) | <0.001 |
| EDUCATION LEVEL  No qualification  School education  Tertiary education  YEARS IN EDUCATION | 100 (15.4)  327 (50.5)  221 (34.1)  12.87 (4.08) | 40 (4)  416 (41.5)  546 (54.5)  14.69 (4.19) | Χ2(2)=102.87  U=8.26 | V=0.25 (0.20-0.3)  r=0.20 | <0.001  <0.001 |
| SOCIAL FUNCTIONING  Employment status  Employed  Unemployed  Marital status  Steady relationship  No relationship  Living arrangements  Independent living  No independent living | 256 (50.3)  253 (49.7)    201 (33.5)  399 (66.5)    220 (42.5)  298 (57.5) | 615 (61.6)  383 (38.4)    626 (62.4)  378 (37.7)    683 (68.5)  314 (31.5) | Χ2(1)=17.7      Χ2(1)=125.2      Χ2(1)=95.96 | V=0.11 (0.06-0.16)  V=0.28 (0.23-0.33)  V=0.25 (0.2-0.3) | <0.001      <0.001      <0.001 |
| OPCRIT DSM IV DIAGNOSIS  Schizophrenia-spectrum disorder  Affective psychosis  Bipolar disorder  Psychotic depression | 409 (71.4)  164 (28.4)  73 (12.7)  91 (15.8) | -  -  -  - |  |  |  |

*SD: standard deviation; df: degrees of freedom*

*2.1.2 eTable 3. Affective vs Schizophrenia-spectrum disorder sociodemographic comparison in white subsample (n=573)*

| **DESCRIPTIVE AT BASELINE** | Number (%)/Mean(SD) | | Statistics |  |  |
| --- | --- | --- | --- | --- | --- |
| Gender  Male  Female   Age (years), mean (SD) | Affective psychosis  n= 164    83 (50.6)  81 (49.4)  32.84 (11.56) | Schizophrenia-spectrum disorder  n= 409    278 (68)  131 (32)  31.63 (10.92) | Tests (df)    Χ2(1)=15.14       z=-1.013 | Effect sizes  V=0.16 (0.09-0.25)  r= -0.042 | p value    <0.001       0.240 |
| EVER USED CANNABIS  No  Yes | 58 (36)  103 (64) | 136 (34.2)  262 (65.8) | Χ2(1)=0.17 | V=0.018 (0.04-0.1) | 0.677 |
| EDUCATION LEVEL  No qualification  School education  Tertiary education  YEARS IN EDUCATION | 25 (15.3)  87 (53.4)  51 (31.3)  12.58 (3.84) | 65 (16.1)  197 (48.6)  143 (35.3)  12.94 (4.12) | Χ2(2)=1.107  Z=0.55 | V=0.04 (0.06-1.13)  r: 0.023 | 0.575  0.581 |
| SOCIAL FUNCTIONING  Employment status  Employed  Unemployed  Marital status  Steady relationship  No relationship  Living arrangements  Independent living  No independent living | 79 (58.5)  56 (41.5)    74 (48.1)  80 (52)    73 (53.7)  63 (46.3) | 141 (45.5)  169 (54.5)    105 (28.3)  266 (71.7)    119 (37.5)  198 (62.5) | Χ2(1)=6.39      Χ2(1)=18.89      Χ2(1)=10.15 | V=0.12 (0.05-0.22)  V=0.19 (0.12-0.28)  V=0.15 (0.08-0.25) | 0.011      <0.001      0.001 |

*SD: standard deviation; df: degrees of freedom*

***2.2 Case-control variance explained of employed PRSs (eFigure1)***

*2.2.1 eFigure1. Variance expressed by Nagelkerke R Square of PRS SZ, BD, D and IQ at 10 different p-values thresholds on case-control (all psychosis vs control) associations adjusted by 10PCs*

PRS D - PGC2 + Biobank

Nagelkerke R Square

|  | Nagelkerke R Square |
| --- | --- |
| p 0.00000005 | 0.000773072 |
| p 0.000001 | 0.0029873 |
| p 0.0001 | 0.00720022 |
| p 0.001 | 0.0133071 |
| p 0.01 | 0.0137548 |
| p 0.05 | 0.0154685 |
| p 0.1 | 0.012578 |
| p 0.2 | 0.0146713 |
| p 0.5 | 0.0123334 |
| p 1 | 0.0130653 |

PRS IQ - Savage 2018

Nagelkerke R Square

|  | Nagelkerke R Square |
| --- | --- |
| p 0.00000005 | 0.00182245 |
| p 0.000001 | 0.00350895 |
| p 0.0001 | 0.00798458 |
| p 0.001 | 0.00835082 |
| p 0.01 | 0.00703113 |
| p 0.05 | 0.0102262 |
| p 0.1 | 0.0141685 |
| p 0.2 | 0.0138322 |
| p 0.5 | 0.0116484 |
| p 1 | 0.0120504 |

PRS BD - PGC2

Nagelkerke R Square

|  | Nagelkerke R Square |
| --- | --- |
| p 0.00000005 | 0.00369574 |
| p 0.000001 | 0.00137034 |
| p 0.0001 | 0.00229917 |
| p 0.001 | 0.00865604 |
| p 0.01 | 0.0236652 |
| p 0.05 | 0.0364056 |
| p 0.1 | 0.0342762 |
| p 0.2 | 0.0289399 |
| p 0.5 | 0.0292451 |
| p 1 | 0.0288994 |

PRS SZ - PGC2

Nagelkerke R Square

|  | Nagelkerke R Square |
| --- | --- |
| p 0.00000005 | 0.00824469 |
| p 0.000001 | 0.0258999 |
| p 0.0001 | 0.0343592 |
| p 0.001 | 0.0481074 |
| p 0.01 | 0.0612091 |
| p 0.05 | 0.0750646 |
| p 0.1 | 0.0819428 |
| p 0.2 | 0.0853845 |
| p 0.5 | 0.0827063 |
| p 1 | 0.0820183 |

***2.3 Visual representation of data***

*2.3.1 eFigure2. Three-dimensional scatterplot of the PRS distribution in the three groups of affective, non-affective psychosis (SSD) and controls*

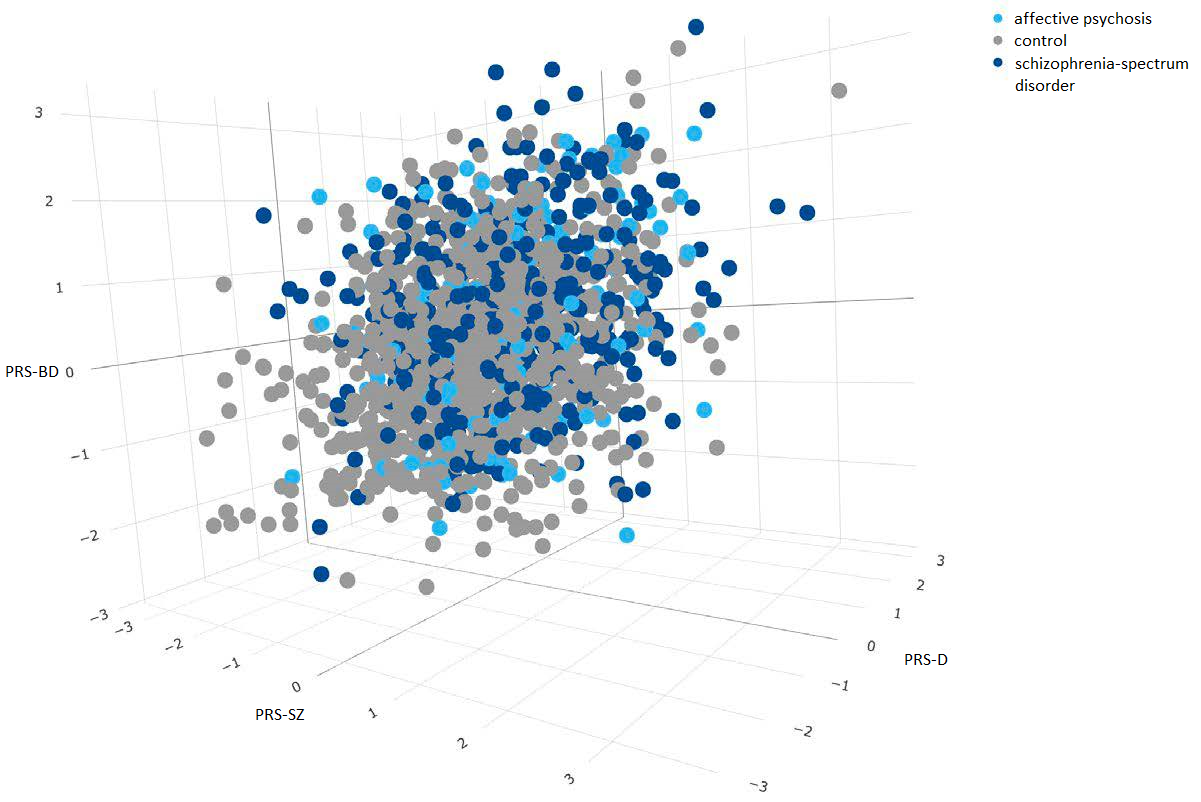

Three-dimensional scatterplot of the PRS distribution in the three groups of affective (AP), non-affective psychosis (SSD) and controls. The three axes correspond to each PRS (z-score after adjustment for PCs and site) and the dots are coloured by group. We observed a large overlap of PRS between the three groups of affective, non-affective psychosis and controls.

***2.4. Detailed results of association of different associations on Model 1 and Model 2.***

*2.4.1 eTable 4. Model 1: Association of different PRSs (SZ, BD, D and IQ) with multicategorical DSMIV* *OPCRIT clinical groups adjusted with 10 PCs in white population (total n=1576)*

| ***Model 1a*** | **PseudoR** | **Prob > chi2** |  |  |
| --- | --- | --- | --- | --- |
| N=1578 | 0.1089 | <0.001 |  |  |
|  | **OR** | **p value** | **95% CI** |  |
| **SSD vs CONTROL** | |  |  |  |
| PRS SZ | **1.87** | **<0.001** | 1.57 | 2.2 |
| PRS BD | **1.34** | **<0.001** | 1.15 | 1.57 |
| PRS D | 1.04 | 0.566 | 0.91 | 1.19 |
| PRS IQ | **0.88** | **0.056** | 0.77 | 1.00 |
| **AP vs CONTROL** | |  |  |  |
| PRS SZ | **1.34** | **0.014** | 1.06 | 1.68 |
| PRS BD | **1.35** | **0.006** | 1.09 | 1.67 |
| PRS D | **1.37** | **0.001** | 1.14 | 1.64 |
| PRS IQ | 0.85 | 0.074 | 0.71 | 1.02 |
| ***Model 1b*** | **PseudoR** | **Prob > chi2** |  |  |
| N=573 | 0.0858 | 0.0013 |  |  |
| **AP vs SSD** | |  |  |  |
| PRS SZ | **0.7** | **0.010** | 0.54 | 0.92 |
| PRS BD | 1.02 | 0.857 | 0.81 | 1.3 |
| PRS D | **1.31** | **0.011** | 1.06 | 1.61 |
| PRS IQ | 0.99 | 0.979 | 0.81 | 1.23 |

*SSD: schizophrenia-spectrum disorder; AP: affective psychosis, SZ: schizophrenia; BD: bipolar disorder; D: depression; IQ: intelligence quotient*

*2.4.2 eTable 5. Model 2: Association of different PRSs (SZ, BD, D and IQ) with multicategorical DSMIV OPCRIT variable adjusted with 10 PCs in white population (non-affective psychosis as reference)*

| ***Model 2a*** | **PseudoR** | **Prob > chi2** |  |  |
| --- | --- | --- | --- | --- |
| N=573 | 0.1106 | 0.0008 |  |  |
|  | **OR** | **p value** | **95% CI** |  |
| **BD vs SSD** |  |  |  |  |
| PRS SZ | 0.97 | 0.865 | 0.68 | 1.39 |
| PRS BD | .98 | 0.893 | 0.71 | 1.35 |
| PRS D | 1.14 | 0.364 | 0.86 | 1.41 |
| PRS IQ | 1.07 | 0.315 | 0.81 | 1.41 |
| **MDD-P vs SSD** |  |  |  |  |
| PRS SZ | **0.52** | **<0.001** | 0.37 | 0.74 |
| PRS BD | 1.04 | 0.814 | 0.77 | 1.4 |
| PRS D | **1.49** | **0.003** | 1.14 | 1.94 |
| PRS IQ | 0.94 | 0.655 | 0.72 | 1.23 |
| ***Model 2b*** | **PseudoR** | **Prob > chi2** |  |  |
| N=164 | 0.20 | 0.0347 |  |  |
| **BD vs MDD-P** |  |  |  |  |
| PRS SZ | **2.14** | **0.007** | 1.23 | 3.74 |
| PRS BD | 1.01 | 0.959 | 0.64 | 1.61 |
| PRS D | 0.71 | 0.092 | 0.48 | 1.06 |
| PRS IQ | 1.03 | 0.878 | 0.71 | 1.49 |

*SSD: schizophrenia-spectrum disorder; AP: affective psychosis, SZ: schizophrenia; BD: bipolar disorder; D: depression; IQ: intelligence quotient*

***2.5 PRS performance for identifying all psychotic diagnostic categories based on OPCRIT using control as reference (eTable 6 and eFigure3).***

We further wanted to study the association of the three four PRS among individual diagnostic categories from the whole psychosis spectrum. Among three PRSs, PRS-SZ presented significant association with most of the diagnostic groups showing the following gradient: SAD (OR=2.38, 95% 1.36 – 4.17, p=0.002) >SZ (OR=2.02, 95% 1.66 – 2.46, p<0.001) >BD (OR=1.76, 95% 1.26 – 2.46, p<0.001) >other psychosis (OR=1.47, 95% 1.1 – 1.98, p=0.009), but not being significantly associated with PD.

Moreover, PRS-BD was significantly associated primarily with other psychosis (OR=1.55, 95% 1.18 – 2.03, p=0.001) and interestingly, PRS-BD showed also a trend of association with MDD-P versus control (OR=1.32 95% CI 1.01 - 1.73, p=0.049).

PRS-D was only significantly associated with MDD-P versus control (OR=1.5, 95% 1.19 – 1.9, p=0.001), and we could not find significant association for PRS-IQ with any group category.).

***eTable 6****. Association of PRSs (for SZ, BD, D and IQ) between OPCRIT diagnostic categories (SZ, OP, SAD, BD and MDD-P) and control as reference adjusted by 10 PCs in white population.*

| ***Model 1*** | **PseudoR** | **Prob > chi2** |  |  |
| --- | --- | --- | --- | --- |
|  | 0.1192 | <0.001 |  |  |
|  | **OR** | **p value** | **95% CI** |  |
| **SCHIZOPHRENIA** | |  |  |  |
| PRS SZ | **2.02** | **<0.001** | 1.66 | 2.46 |
| PRS BD | **1.31** | **0.003** | 1.09 | 1.56 |
| PRS D | 1.00 | 0.962 | 0.86 | 1.17 |
| PRS IQ | 0.90 | 0.165 | 0.77 | 1.05 |
| **OTHER PSYCHOSIS** | |  |  |  |
| PRS SZ | **1.47** | **0.009** | 1.10 | 1.98 |
| PRS BD | **1.55** | **0.001** | 1.18 | 2.03 |
| PRS D | 1.20 | 0.114 | 0.96 | 1.52 |
| PRS IQ | 0.87 | 0.241 | 0.69 | 1.10 |
| **SCHIZOAFFECTIVE DISORDER** | |  |  |  |
| PRS SZ | **2.38** | **0.002** | 1.36 | 4.17 |
| PRS BD | 1.08 | 0.770 | 0.64 | 1.83 |
| PRS D | 0.85 | 0.448 | 0.56 | 1.29 |
| PRS IQ | 0.71 | 0.130 | 0.46 | 1.10 |
| **BIPOLAR DISORDER** | |  |  |  |
| PRS SZ | **1.76** | **<0.001** | 1.26 | 2.46 |
| PRS BD | 1.34 | 0.057 | 0.99 | 1.82 |
| PRS D | 1.21 | 0.150 | 0.93 | 1.58 |
| PRS IQ | 0.91 | 0.510 | 0.70 | 1.19 |
| **PSYCHOTIC DEPRESSION** | |  |  |  |
| PRS SZ | 1.07 | 0.651 | 0.80 | 1.44 |
| PRS BD | **1.32** | **0.049** | 1.00 | 1.73 |
| PRS D | **1.50** | **0.001** | 1.19 | 1.90 |
| PRS IQ | 0.80 | 0.063 | 0.64 | 1.01 |

*SZ: schizophrenia; BD: bipolar disorder; D: depression; IQ: intelligence quotient*

*SZ (n=487); Other psychosis (n=205); Schizoaffective disorder (n=47); BD (n=130), MDD-P (n=137)*

***eFigure 3.*** *PRS performance across all psychotic spectrum DSM4 OPCRIT categories.*

**p<0.05*

***p<0.001*

PRS distribution across OPCRIT diagnosis

OR

|  | PRS SZ | PRS BD | PRS D | PRS IQ |
| --- | --- | --- | --- | --- |
| Schizophrenia | 2.02 | 1.31 | 1 | 0.9 |
| Other psychosis | 1.47 | 1.55 | 1.2 | 0.87 |
| Schizoaffective Disorder | 2.38 | 1.08 | 0.85 | 0.71 |
| Bipolar Disorder | 1.76 | 1.34 | 1.21 | 0.91 |
| Psychotic Depression | 1.07 | 1.32 | 1.5 | 0.8 |

*SZ: schizophrenia; BD: bipolar disorder; D: depression; IQ: intelligence quotient*

**2.6 *Goodness of fit of data of join model combining three major psychiatric disorder polygenic scores (SZ, BD, MDD-P) and polygenic score for intelligence for SSD and AP comparison.***

***eFigure4.***

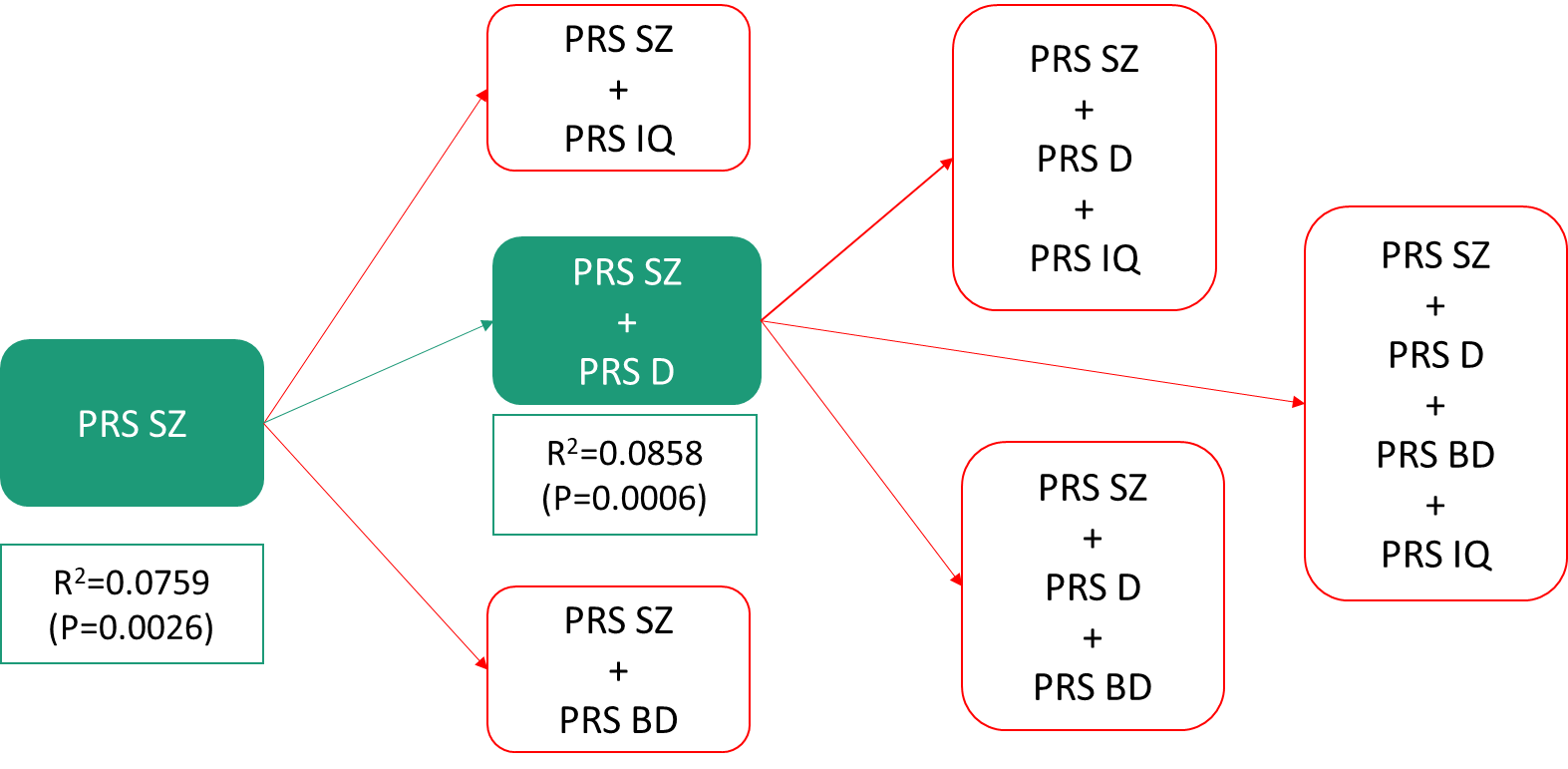

*Green lines represent improvement of model. Red lines represent non-significant likelihood-ratio tests. SZ: schizophrenia; BD: bipolar disorder; D: depression; IQ: intelligence quotient*

Goodness of fit of data was explored through likelihood ratio test while sequentially adding the four PRSs to the models in order to identify those PRS adding value to the discriminability between clinical groups (SSD and AP). The best fitness of data by per likelihood ratio test was by adding PRS-SZ and PRS-D to the model (Δχ^2^(1) = 6.74, p = 0.0094).

**eAppendix 3. References**

1	Mallett, R., Leff, J., Bhugra, D., Pang, D. & Zhao, JH. Social environment, ethnicity and schizophrenia. *Soc Psychiatry Psychiatr Epidemiol*; **37**: 329–335 (2002).

2	Purcell, S. *et al.* PLINK: A tool set for whole-genome association and population-based linkage analyses. *Am J Hum Genet*; **81**: 559–575 (2007).

3	Price, AL. *et al.* Long-Range LD Can Confound Genome Scans in Admixed Populations. Am. J. Hum. Genet. ; **83**: 132–135 (2008).

4	Ripke, S., Neale, BM., Corvin, A., Walters, JT. & Schizophrenia Working Group of the Psychiatric Genomics Consortium. Biological insights from 108 schizophrenia-associated genetic loci. *Nature*; **511**: 421–427 (2014).

5	Stahl, EA. *et al.* Genome-wide association study identifies 30 loci associated with bipolar disorder. *Nat Genet*; **51**: 793–803 (2019).

6	Howard, DM. *et al.* Genome-wide meta-analysis of depression identifies 102 independent variants and highlights the importance of the prefrontal brain regions. *Nat Neurosci*; **22**: 343–352 (2019).

7	Savage, JE. *et al.* Genome-wide association meta-analysis in 269,867 individuals identifies new genetic and functional links to intelligence. *Nat Genet*; **50**: 912–919 (2018).

8	American Psychiatric Association. *Diagnostic and statistical manual of mental disorders : DSM-IV.* 4th ed., T. American Psychiatric Association: Washington, DC, 1994.

9	Morgan, C. *et al.* Modelling the interplay between childhood and adult adversity in pathways to psychosis: initial evidence from the AESOP study. *Psychol Med*; **44**: 407–19 (2014).

10	Hair, J., Black, W., Babin, B. & Anderson, R. *Multivariate Data Analysis*. 7th Editio. 2014www.pearsoned.co.uk (accessed 29 Apr2020).

11	Dudbridge, F. Power and Predictive Accuracy of Polygenic Risk Scores. *PLoS Genet*; **9**: e1003348 (2013).

12	Ripke, S. *et al.* Biological insights from 108 schizophrenia-associated genetic loci. *Nature*; **511**: 421–427 (2014).
